# Patient knowledge, attitudes and practices on chronic wound infections in Tanga Regional Referral Hospital, Tanzania; a qualitative study

**DOI:** 10.1101/2025.05.12.25327278

**Authors:** Aleena Dawer, Victor Msengi, Pendo Magili, John P.A. Lusingu

## Abstract

**Background:** Bacterial wound infections contribute significantly to global mortality, with nearly half of contaminated wounds progressing to chronic states, which can result in amputations. In Tanzania, the situation is worsened by the misuse of antibiotics and the rise of drug-resistant bacteria, alongside patients’ varied understanding of wound care management. This qualitative study explores patient knowledge, attitudes, and practices (KAP) regarding chronic wound infections in Tanzania.

**Methods:** A sample of 15 patients was selected from both genders, representing a wide range of ages and wound severities. Semi-structured interviews were conducted in Swahili at the Tanga Regional Referral Hospital (TRRH) from October 2023 to April 2024.. Interviews used open-ended questions on dietary habits, traditional medicine, medication adherence, and wound care practices. Social demographic data were collected to contextualize patient experiences. Interviews were audio-recorded, transcribed verbatim, and analyzed via Nvivo.

**Results:** Thematic analysis of the interviews identified six major themes: delays in treatment-seeking, inadequate wound management, reliance on ritual practices, poor eating behaviors, challenges in coping with chronic wounds, and inadequate healthcare services. Delays stemmed from misjudging wound severity and reliance on traditional healers. Financial constraints contributed to inadequate wound management, with many patients turning to herbal remedies. Nutritious food access was limited, primarily due to financial and availability constraints. Patients faced substantial physical, emotional, and financial burdens. Long wait times and inconsistent care were also major barriers to effective wound management.

**Conclusion:** Chronic wound infections are exacerbated by systemic healthcare shortcomings and limited patient education. Insights from patient perspectives emphasizes the need for proper wound management, understanding of antimicrobial resistance (AMR), and addressing cultural beliefs and resource constraints. Patient-centered models can foster more effective and sustainable models of wound care in the community setting.

## Introduction

Chronic wound infections are a pervasive global health issue, significantly diminishing patient quality of life and placing a substantial burden on healthcare systems. Defined as wounds that fail to heal within three months, chronic wounds often persist due to a complex interplay of biological, socioeconomic, and healthcare-related factors. These infections are a leading cause of major lower-limb amputations, with systematic review studies showing that up to 50% of patients die within five years of undergoing such procedures^1^. Diabetic foot ulcers (DFUs) alone account for approximately 85% of these amputations worldwide^2^. Yet many of these outcomes are preventable with timely diagnosis and effective treatment.

Understanding the patient perspective is essential to effectively address the challenges of chronic wound management. Current research highlights a significant deficiency in patient understanding about proper wound care and the appropriate use of antibiotics, leading to suboptimal treatment outcomes and increased risk of AMR. For instance, 20% of Europeans reported using antibiotics to self-treat influenza, despite antibiotics being ineffective against viral infections^3^. Similarly, in Kenya, a detailed analysis in selected hospitals in Kajiado County found correlations between septic chronic wounds with individual understanding, compliance with prescribed drug dosages, alcohol consumption, and smoking habits^4^. In the U.S., a study found that 38% of patients were unaware of proper wound dressing techniques and 58.7% lacked education on wound cleansing after hospital discharge^5^. These issues are magnified in resource-limited settings, where patient education and healthcare access are often inadequate.

In Tanzania, where antibiotic misuse is common and healthcare infrastructure is stretched, managing chronic wound infections poses unique challenges. A regional overview of antimicrobial resistance (AMR) in Tanga, a coastal region in northeastern Tanzania, is provided in the quantitative analysis of this study^6^. Despite the clinical importance of managing chronic wounds, there is a lack of published research exploring how Tanzanian patients perceive and manage wound infections during hospitalization. This study addresses that gap by conducting in-depth, semi-structured interviews with hospitalized patients in Tanga Regional Referral Hospital.

## Methods

### Study aim, design and selection criteria

This study employed a cross-sectional approach conducted from 27/10/2023-01/04/2024 to explore the experiences, challenges, and perceptions of patients suffering from chronic wound infections at the Tanga Regional Referral Hospital (TRRH). The selection criteria focused on all admitted patients of all ages and sexes exhibiting chronic wound infections. These criteria were chosen to provide comprehensive insights into the factors affecting chronic wound management in this clinical setting.

### Sample size and population

The focal point of this study was the diverse group of patients admitted to TRRH with chronic wounds. This population includes both males and females who have been afflicted with chronic wounds. 15 patients were selected for in-depth interviews (IDIs), ensuring a mix of age, gender, and wound severity to capture diverse experiences. The chosen number of patients was believed to be sufficient to reach thematic saturation based on consultation with a qualitative expert. As interviews progressed, no new themes emerged, indicating that key patterns were captured and saturation was reached

### Study setting

TRRH, commonly referred to as Bombo Regional Hospital, is a prominent hospital located in Tanga, Tanzania. Established during the German colonial period in the 1890s to combat high malaria mortality rates, TRRH has since evolved into a key medical facility. The hospital, which admits an average of 28 patients per month for chronic wound infections, serves as a crucial center for both routine and specialized healthcare services in the region.

### Sampling strategy

A consecutive sampling strategy was adopted for this study. Between 27/10/2023-01/04/2024, all patients meeting the inclusion criteria were approached for participation until the desired sample size was achieved.

### Qualitative method

Participants had variability in age, gender, wound severity, and treatment delays, which offered distinct insights into patient experiences. Individual interviews were chosen over focus groups to encourage candid and uninfluenced accounts of patient experiences with chronic wounds and healthcare interactions.

Participants were thoroughly informed about the study’s objectives and procedures, and informed consent was obtained from each participant. The interviews, designed to last between 10 to 15 minutes, were conducted using open-ended questions in Swahili to reduce language and cultural barriers and facilitate a more natural dialogue. The interviews followed a structured guide divided into two sections. The first section explored dietary habits, use of traditional medicine, medication adherence, and wound care practices. The second section focused on challenges faced by patients, their suggestions for improvement, and their perspectives on their condition and treatment.

Sociodemographic data, including age, gender, residence, education level, occupation, religion, and wound type and duration, were collected to provide a comprehensive understanding of patient experiences. The study explored both practical treatment aspects and personal, subjective perspectives.

A significant emphasis was placed on the comfort and privacy of participants during interviews. Participants were allowed to choose their preferred location for the interview, ensuring that their perspectives and experiences were shared freely and without interference. Data recording involved the use of audio recorders and note-taking. Post-interview, the audio-recorded interviews were transcribed verbatim and translated from Swahili into English. The transcripts were carefully reviewed alongside the audio recordings to ensure accuracy and fidelity to participants’ responses.

### Data analysis

Thematic analysis was conducted using NVivo software, following an iterative process. An initial codebook, outlining deductive codes, was developed based on the study’s predefined objectives. Two researchers independently coded a subset of transcripts to refine the codebook, resolve discrepancies, and ensure reliability.

Four main deductive themes were confirmed during coding, while inductive analysis revealed two additional emergent themes. Constant comparison across transcripts ensured thematic saturation. NVivo’s coding queries and matrix searches systematically grouped excerpts and explored relationships between themes and participant characteristics.

Coded data were synthesized into overarching categories and sub-themes, supported by illustrative quotations. Memo-writing was used throughout the process to document analytic decisions and track evolving interpretations. This rigorous, multi-step approach ensured that the final thematic structure provided a credible representation of participants’ experiences with chronic wound management.

## Results

Fifteen patients (age range: 35-80 years; median: 55 years) participated, with 93% presenting leg wounds. Gender distribution was relatively balanced, with eight males and seven females. Participants were fairly evenly split between Christian (53.3%) and Muslim (46.7%) faiths. Educational attainment was predominantly limited to primary school education (73.3%), with fewer participants having achieved secondary education (13.3%) or college-level education (6.7%). Nearly half (46.7%) worked in agriculture, which is consistent with regional workforce patterns (47.5% of women and 51.9% of men in Tanga work in farming)^7^.

The sample’s older median age (55 vs. Tanga’s ∼25 years) reflects chronic wounds’ association with aging^8^.. Educational attainment among participants was consistent with regional averages, where adults’ mean years of education is approximately six years^7^. The religious distribution maintained regional representativeness.

Table 2 outlines major themes and their related codes from patient interviews, covering areas such as delays in seeking treatment, wound management, ritual practices, eating behaviors, coping with chronic wounds, and challenges with healthcare services. These themes capture various factors affecting patient care and experience at TRRH. The analysis initially identified four themes, with two additional inductive themes—“Living and Coping with Chronic Wounds” and “Inadequate Healthcare Services”. As the interviews progressed, no significant new themes emerged. Participants’ responses consistently reiterated previously identified ideas, indicating that key patterns in the data had been captured and that data saturation had been reached.

**Table 1.**
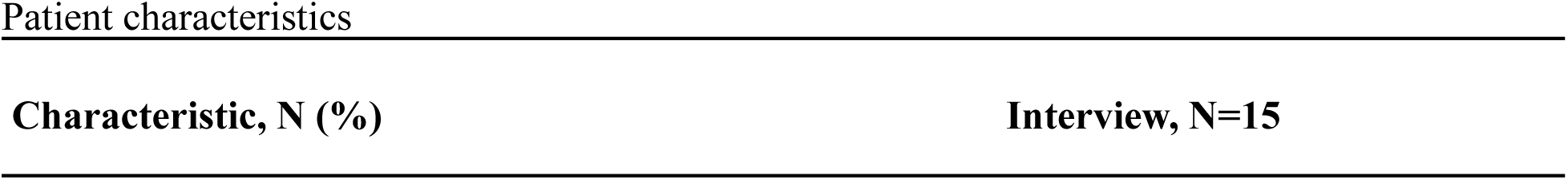
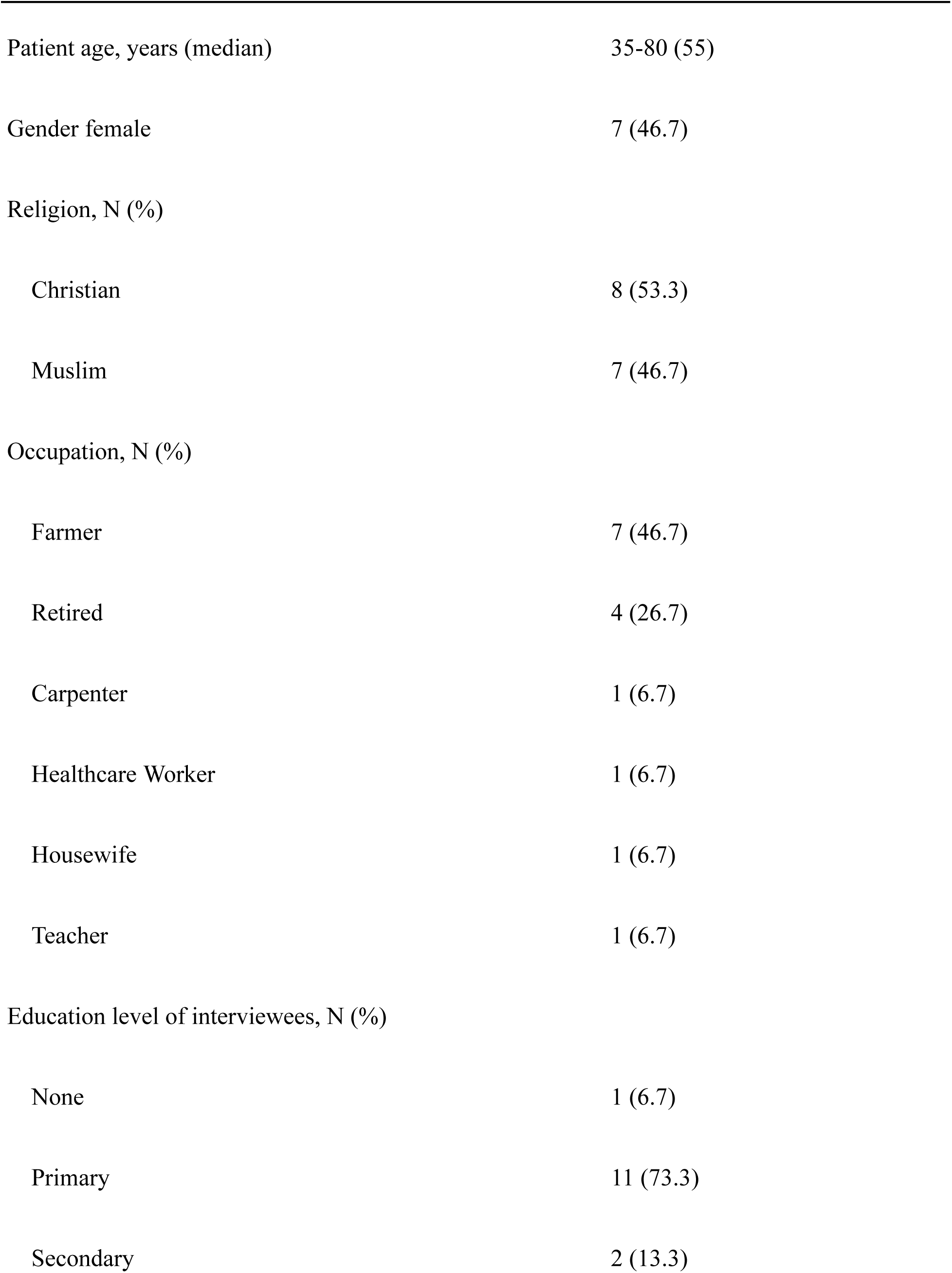
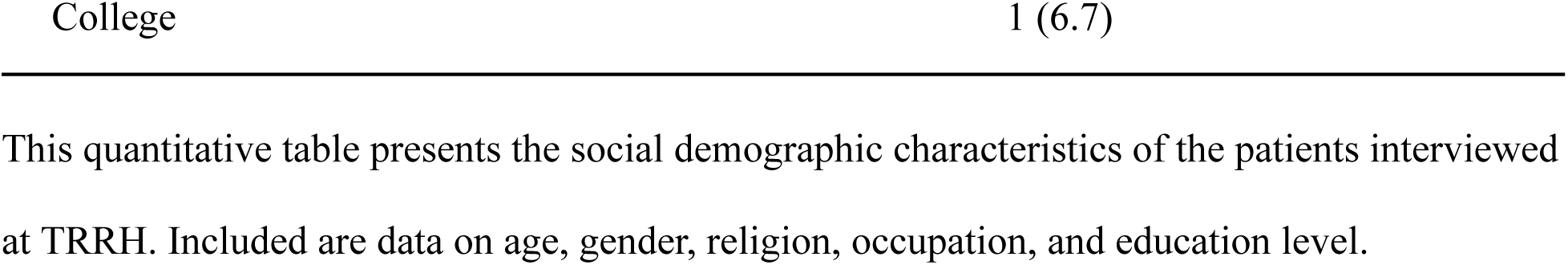
Demographic characteristics of TRRH patients interviewed.

**Table 2:**
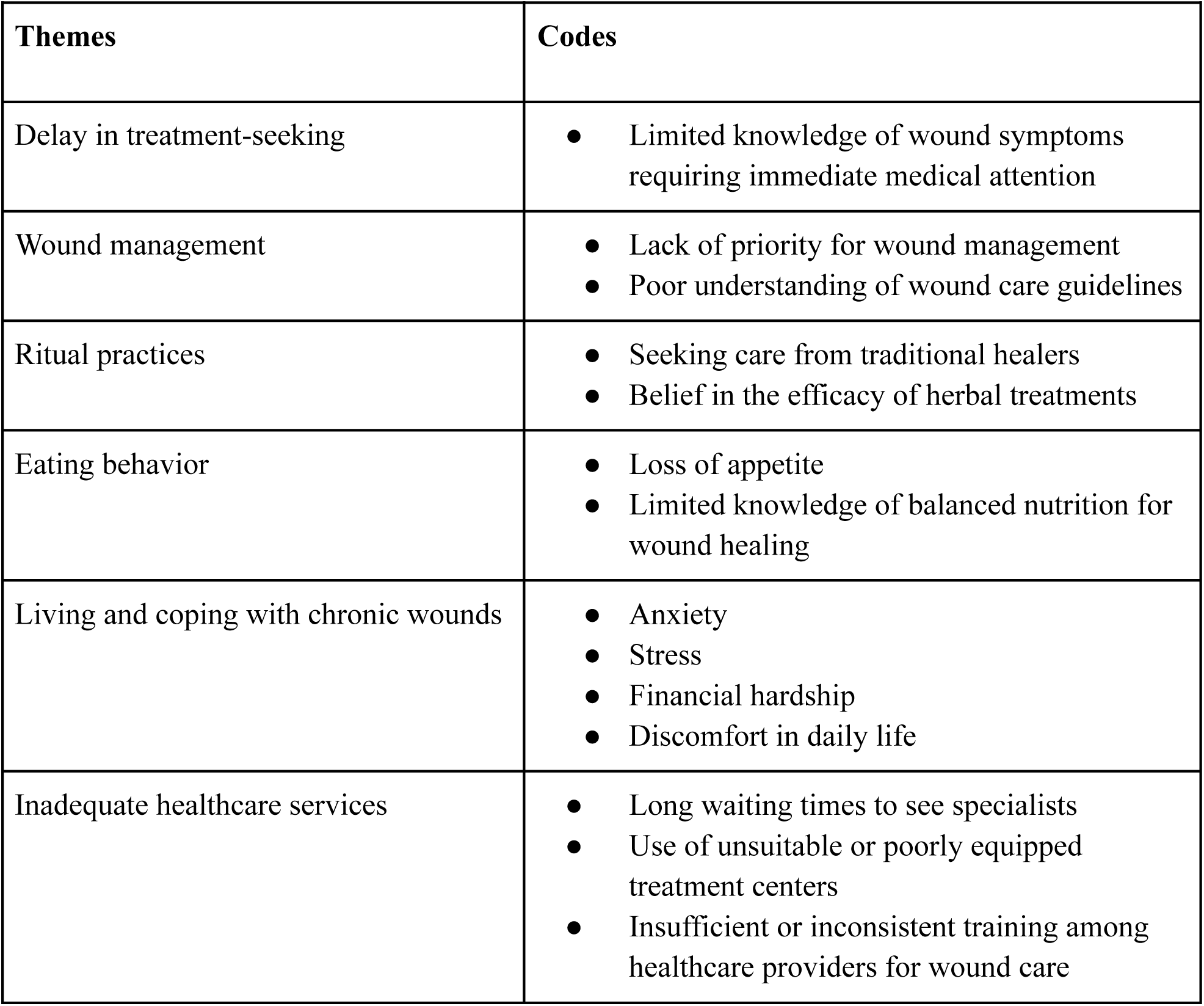
Summary of themes and codes.

| Themes | Codes |
| --- | --- |
| Delay in treatment-seeking | • Limited knowledge of wound symptoms requiring immediate medical attention |
| Wound management | • Lack of priority for wound management<br>• Poor understanding of wound care guidelines |
| Ritual practices | • Seeking care from traditional healers<br>• Belief in the efficacy of herbal treatments |
| Eating behavior | • Loss of appetite<br>• Limited knowledge of balanced nutrition for wound healing |
| Living and coping with chronic wounds | • Anxiety<br>• Stress<br>• Financial hardship<br>• Discomfort in daily life |
| Inadequate healthcare services | • Long waiting times to see specialists<br>• Use of unsuitable or poorly equipped treatment centers<br>• Insufficient or inconsistent training among healthcare providers for wound care |

### Theme 1: Delay in treatment-seeking

Postponed hospital treatment was a significant theme among patients with wound infections. Most patients expressed a lack of understanding of severe wound symptoms, leading them to attempt self-care or seek help from traditional healers. Hospital care was often only sought once the wound had significantly worsened. The findings here indicate an urgent need for enhancing public awareness about recognizing wound symptoms early and understanding the risks of delayed medical care, including the potential for severe infections and the spread of antibiotic-resistant strains.

> *“It took like six months without going to the hospital…” (P2, Male)*
>
> *“Unfortunately, I thought it was a fungus [and] treated it with antifungal drugs. Only when my leg became swollen and turned brown, then I knew there was a problem…” (P9, Male)*
>
> *“I stayed at home for a while and went to the pharmacy after seeing that my leg was swollen. I got five injections for five days without success, so I went to the hospital. After arriving they said, “the leg is already in a bad condition” (P13, Male)*

### Theme 2: Wound management

Participants described their methods of wound care both before and after hospital visits. Their responses revealed limited familiarity with formal wound care guidelines. Many reported using readily available materials, such as non-sterile cotton swabs or pieces of cloth, to clean and cover their wounds. For example, P1 described cleaning and covering the wound with a cloth. While participants often used whatever materials were available, using non-sterile coverings raises concerns about potential infection risks.

Financial constraints also played a critical role in hindering adherence to recommended wound care practices. Participants like P4 recounted how financial constraints forced them to skip necessary dressing changes at the clinic, highlighting the harsh reality of having to choose between proper medical care and other daily expenses. Such decisions not only compromise wound healing but also escalate the risk of complications. P5 illustrated the dire consequences of inadequate wound care in the context of chronic conditions like diabetes, which can lead to severe outcomes such as amputations.

> *“Ah, for the first month I cleaned it every day, but in the following month, I would skip a day and then go dress it… I was unable to go frequently because of the financial challenge… for dressing and transport cost to reach the health center every day…” (P4, Female)*

### Theme 3: Ritual practices

Patients frequently turned to traditional healers and plant-based remedies for managing chronic wounds, often attributing their conditions to supernatural causes like witchcraft. This theme is critical as it reveals the cultural belief systems influencing health-seeking behaviors for chronic wounds.

Participants often preferred traditional healers over medical facilities, as exemplified by P2, who visited eight healers over three years in search of effective remedies. Despite his efforts, the treatments worsened his condition, highlighting the desperation and hope that drove his reliance on these practices. The diverse explanations for his wound given by different healers, ranging from mystical interpretations like “two birds with two beaks” causing the wound or having “offended mermaid children,” reflect the varied and often complex belief systems surrounding illness in Tanzania.

The use of plant-based remedies further illustrates the reliance on traditional healing practices. For instance, P6 used *Mzambarao* bark (*Java Plum* or *Syzygium cumini*), recommended by another diabetic patient, thought to reduce symptoms like frequent urination. Similarly, P9 applied a leaf to his wound after dissatisfaction with hospital medicines, trusting in natural treatments perceived as safer and more effective than conventional care.

Participants’ reliance on traditional healers and herbal remedies not only demonstrates a cultural inclination but also possibly reflects underlying issues in the healthcare system, such as accessibility, affordability, and trust. The persistence of these practices, despite instances of adverse effects, underscores the profound influence of cultural and traditional beliefs in health management, especially in communities where such practices are deeply ingrained.

While some traditional practices delayed access to conventional medical treatment, certain plant-based remedies used by participants may hold genuine therapeutic value. For instance, Mzambarao bark has been studied for its potential antidiabetic and anti-inflammatory properties.^9^ The use of herbal treatments reflects not just cultural preferences but also historical knowledge of local medicinal plants. Integrating evidence-based understanding of traditional remedies into healthcare could improve delivery by fostering trust and offering more culturally attuned care.

> *“I visited like eight traditional healers, used traditional medicine for three years… the condition worsened … you apply medicine, the leg becomes sore and itchy…” (P2, Male)*
>
> *“No, I don’t use any herbal medicines” (P4, Female)*
>
> *“The second [healer] spoke of two birds with two beaks, saying that’s why I have sores here and there, that’s what’s troubling you. (Laughter)…the third claimed spirits wanted me to become a traditional healer…another said I stepped on mermaid children…at a crossroads…” (P2, Male).*
>
> *“I used the bark of a tree called Mzambarao… effectiveness was…equivalent to the pills because it lowered the sugar level faster…” (P6, male).*
>
> *“Every time I take these hospital medicines I don’t get relief, I met a gentleman, he said let me give you medicine. He gave me a leaf and I chewed it and put it on my wound.” (P9, Male)*

### Theme 4: Eating behavior

Participants’ accounts revealed that economic hardship and illness-related appetite loss were major barriers to maintaining diets that support wound healing. Some participants acknowledged the importance of food in recovery but faced significant challenges in accessing adequate nutrition. Protein-rich foods such as meat were described as unaffordable, and several participants reported skipping meals due to financial constraints.

For instance, P2 mentioned eating meat rarely due to financial constraints, P4 skips lunch, while P5 described relying on porridge and struggling with blood sugar stabilization despite eating minimally.

> *“I have never followed any diet, I eat ugali, rice and sometimes miss meals completely…I eat meat but once in a while depending on the economic situation, if I get it today I might go a whole month without it again.” (P2, Male).*
>
> *“At night… we eat light foods like bananas… or cassava with tea.” (P1, Female)*
>
> *“I often don’t eat at lunch.” (P4, Female)*
>
> *“In the morning, I can only drink porridge…And eat nothing until the next morning…I can’t taste food…But surprisingly, the sugar level doesn’t stabilize.” (P5, Male).*

### Theme 5: Inadequate healthcare services

Poor healthcare services can significantly hinder progress in health improvement, particularly when individuals with serious health conditions, such as chronic wounds, seek treatment and discover that immediate care is unavailable. Participants’ experiences reveal a range of issues from delayed treatment to inadequate guidance on wound care, painting a picture of a healthcare system struggling to meet the needs of patients with complex conditions.

Delays in treatment were a recurring issue, with P9 describing a critical delay upon hospital arrival that worsened his condition, and P10 recounting four days without treatment at a district hospital before being transferred to a referral hospital.

Inadequacies in wound care and management advice were also evident. Several participants, such as P1, P3, P12, and P15, noted a lack of detailed instructions from healthcare workers (HCWs) on cleaning and managing their wounds. This lack of guidance often left patients to rely on their understanding or alternative remedies. P2’s account of his wound worsening despite receiving basic care at a health center highlights the complexity of wound management, where both healthcare guidance and the patient’s ability to follow at-home care practices play critical roles in outcomes.

Participants also reported inconsistencies in treatment protocols across facilities. P4, for instance, described differing approaches, from povidone and metronidazole to honey dressings. This lack of standardization and clear communication undermines patient trust and contributes to poor outcomes, as seen with P5, who ultimately required a toe amputation due to inadequate initial care and follow-up.

> *“Since I arrived here on Tuesday evening, they have already set their treatment time table only for Tuesday and Wednesday of next week. So it bothered me and it was difficult to get treatment and I see that the bacteria are attacking more and more now…” (P9, Male)*
>
> *“I went to the district hospital, I didn’t get any treatment there. I stayed there for four days, so I had to come back to the referral hospital …” (P10, Male)*

### Theme 6: Living and coping with chronic wounds

The testimonies reflect a spectrum of challenges, from physical and emotional struggles to socioeconomic constraints, all of which paint a vivid picture of the multifaceted burden of a chronic wound. Patients frequently expressed profound anxiety, particularly about potential amputations, as seen in P6, who also described the distress caused by foul-smelling discharge affecting his social interactions and self-esteem.

Physical challenges were also prominent, as illustrated by P1, a female, who struggled with swollen feet, making it difficult to walk or wear shoes. This physical limitation represents just one aspect of the broader impact on daily life and mobility. P3 recounted a harrowing experience of unconsciousness and severe burns, highlighting the drastic, life-altering consequences of chronic conditions and accidents, which are compounded when layered with chronic wound management.

Economic constraints were another critical aspect, with patients like P3 discussing the financial burdens of treatment and the reliance on their children to support them. This economic stress can lead to delayed or inadequate treatment, exacerbating wound progression. P2 detailed the challenge of adhering to treatment schedules and the demoralization felt when treatments do not yield immediate results, reflecting the psychological toll of managing a chronic condition.

A chronic wound is not just the physical pain but also the psychological distress, social challenges, and economic burdens faced by patients. All patients (n=15) expressed the emotional toll of anxiety and isolation that ripples into their lives, affecting their relationships.

> *“Ah, challenges, truly…just worries. Anxiety because often with diabetic wounds, you hear about people having their legs amputated…you just feel the pain, it’s like pricking - Ah - it’s sharp. Secondly, the fluid it releases has a bad smell, so when you sit with others… you find flies following you, you can’t sit comfortably.” (P6, Male)*
>
> *“My body becomes very heavy, I can’t handle it…For example, I might go outside, but it’s a challenge. I might fall…It feels like my legs are weakening, yes. And, you might use the toilet but then find it hard to get up. In short, I can’t manage on my own.” (P5, Male).*
>
> *“Ah, I don’t really understand the medications I take.” (P10, Male).*

## Discussion

This study fills a critical gap in understanding patients’ knowledge, attitudes, and practices (KAP) regarding chronic wound infections in Tanzania, an area with limited prior research. While studies in Sub-Saharan Africa (SSA) have examined HCW perspectives on wound care, such as in Ghana^10^, few have focused on patients’ lived experiences, which is essential for improving wound management practices. This study aligns with global findings on wound care while highlighting unique challenges shaped by the sociocultural and healthcare dynamics of a specific region within SSA.

Globally, patient KAP regarding wound infections share commonalities but also reveal regional differences. For example, a Canadian study^11^ reported the significant emotional and psychological toll of chronic wounds, with patients expressing frustration, anxiety, and concerns about delayed healing. Similarly, Tanzanian participants in this study described fear of amputation and the stigma associated with chronic wounds, including embarrassment over foul odors and flies, which impacted their social interactions and mental well-being. Both studies also emphasized the challenge of self-managing symptoms, with pain, fatigue, and difficulty concentrating being debilitating and affecting daily activities. However, a notable difference was that the Canadian study revealed numerous concerns among patients regarding burdening their family members, a sentiment rarely expressed in the Tanzanian context.

### Treatment disparities

Beyond psychosocial experiences, disparities in available treatment options also play a crucial role in shaping patient outcomes in chronic wound management. While parallels exist in the emotional and psychological burdens experienced by patients globally, stark differences in treatment availability highlight inequalities in healthcare infrastructure. Canadian patients had access to advanced interventions such as sharp debridement and compression bandages, whereas Tanzanian participants relied on basic treatments like honey dressings and, in severe cases, amputations. The use of honey as a wound dressing agent at TRRH mirrors findings by previous research^12^ which documented its widespread use in SSA for its natural healing properties. This integration of traditional and allopathic practices underscores the need for context-specific solutions in resource-limited settings.

### Cultural and economic influences on traditional medicine

A predominant theme in this study was the reliance on traditional healing practices, driven by cultural beliefs and economic constraints. Similar to findings in previous studies,^13,14^ patients turned to plant-based remedies due to their perceived effectiveness, affordability, and alignment with cultural norms. However, the lack of standardization in traditional practices, such as inconsistent dosage measurements, raises concerns about safety and efficacy. In a meta-literature review,^15^ it was reported that 80% of Africa’s population relies on medicinal plants as their primary form of medication, a finding reflected in this study, where many patients sought traditional remedies before transitioning to hospital treatments.

Economic factors further shaped healthcare decisions, with limited finances driving reliance on affordable traditional medicine over formal medical care. This aligns with previous observations^16^ that inaccessible healthcare facilities and financial constraints are critical barriers to seeking professional treatment.

While the specific remedies varied by context, reliance on traditional healing practices is not unique to Tanzania. For example, a study in Saudi Arabia^17^ found that cultural practices for wound healing included the use of incense (36.8%), coffee beans (24.3% believing they could stop bleeding), and Sabkha (19% using it as an anti-inflammatory remedy). Such findings highlight that while traditional medicine is widespread across different regions, the specific substances, practices, and cultural interpretations of healing vary.

Despite standardization challenges, emerging evidence highlights the pharmacological potential of many traditional remedies. Ethnobotanical research in sub-Saharan Africa has documented over 2,000 medicinal plants with therapeutic properties, including antimicrobial, anti-inflammatory, and wound-healing effects^18^. A Sri Lankan study similarly reported that 68.1% of patients initially used “hand medicines” for diabetic wounds^19^. With further study and careful integration, these remedies could complement formal healthcare systems, enhancing culturally appropriate care and expanding treatment access in resource-limited settings.

### Dietary practices and nutrition

This study highlighted the major role that financial constraints and food accessibility challenges played in shaping participants’ diets during wound recovery. While many recognized the importance of nutrition for healing, they struggled to afford protein-rich or nutrient-dense foods. Limited financial resources often resulted in inadequate dietary intake, with patients relying on inexpensive staples like porridge, cassava, and bananas and frequently skipping meals altogether. This observation parallels findings from a Colombian HIV study,^20^ where 61.8% of participants faced similar challenges in food selection and purchasing due to financial and informational barriers. For improved recovery outcomes, interventions must combine patient education on nutrition’s role in wound healing with systemic efforts to increase affordability and access to high-protein foods.

### Barriers in healthcare access

Another critical challenge was delayed access to healthcare, a global barrier in managing chronic wounds. Prolonged wait times for treatment, criticized in multidisciplinary wound clinics in Canada^9^, were also present in Tanzania, where participants reported significant delays in receiving care. In Tanzania, delays were often linked to long travel distances to hospitals, limited availability of specialized wound care services, and financial constraints that delayed initial care-seeking. In some cases, inefficiencies in the referral system further compounded these delays, with patients seeking care at multiple facilities before receiving appropriate treatment.

The consequences of delayed care were severe; participants in this study described irreversible damage, including infections worsening to the point of requiring amputations. Similar patterns were observed in a study from Sri Lanka, where 21.7% of patients undergoing delayed care ultimately required amputation^19^. These compounding delays worsen patient outcomes, highlighting the urgent need for structural reforms to ensure timely and equitable wound care access.

### Gaps in patient education

A recurring theme across studies was the limited engagement of HCWs in patient education. One transnational review of leg ulcer treatment adherence^21^ reported that only 50% of nurses provided health education to leg ulcer patients. This trend is echoed in this study, where participants noted insufficient guidance on wound care and lifestyle management. Similarly, misconceptions about wound care were observed in other contexts; for example, in the Saudi study^18^, 41.5% of participants mistakenly believed that showering delayed wound healing, highlighting the importance of medical education and engagement with HCWs.

Such misconceptions, compounded by limited professional guidance, highlight the importance of investing in HCW training and allocating resources to empower patients through education. Globally, research on patient KAP in the context of wound infections has largely been conducted outside of East Africa, limiting its relevance to the region’s unique challenges. By capturing patient perspectives from Tanzania, this study contributes regionally grounded patient insights into chronic wound management; highlighting the need for context-specific interventions and laying a foundation for future research in similar resource-limited settings.

### Limitations

While this study provides valuable insights into wound care challenges in resource-limited settings, several limitations should be noted. This study was conducted with a small sample of patients from a single facility, limiting the findings’ generalizability. We do not claim to represent the full spectrum of wound types, severities, or patient experiences across Tanzania. Instead, the study serves as an exploratory blueprint for future research by offering initial insights into the healthcare-seeking behaviors, coping strategies, and wound management challenges faced by patients in resource-limited settings.

The qualitative approach, while ideal for capturing patient perspectives in depth, requires validation through larger, mixed-methods studies. Interview translation from Swahili to English posed potential accuracy risks, though these were mitigated through collaborative analysis with bilingual, culturally fluent researchers.

These limitations notwithstanding, the study offers foundational knowledge about healthcare-seeking behaviors and coping strategies in Tanzania’s chronic wound population, a previously understudied area.

## Conclusions

This study reveals critical gaps in Tanzania’s chronic wound care landscape, particularly regarding health literacy, healthcare access, and the complex interplay of psychological, cultural, and socioeconomic barriers. Through patient-centered exploration, it identifies actionable themes to guide future interventions for improved outcomes.

While offering an early in-depth exploration of these challenges in the Tanzanian context, our findings underscore the need for three key developments: expanded research to validate these patterns across diverse clinical settings, culturally-adapted wound care protocols that bridge traditional and biomedical approaches, and health system reforms addressing both affordability and care coordination. These exploratory results establish an essential foundation for developing context-specific solutions that address the complex realities of chronic wound management in resource-limited environments, where patient needs intersect with structural healthcare constraints in particularly consequential ways.

## Data Availability

The data underlying this study consist of qualitative interview transcripts that contain sensitive and potentially identifiable patient information. Due to ethical and confidentiality considerations, these data cannot be shared publicly. Researchers interested in accessing de-identified excerpts for academic purposes may contact the corresponding author upon reasonable request and with appropriate ethical approvals.

## Declarations

### Ethics approval and consent to participate

The proposed study is grounded in the ethical principles of beneficence, non-maleficence, and respect for autonomy. The anticipated benefits of the study, which include enhancing knowledge about chronic wound infections and potentially developing more effective treatment strategies, justify the ethical conduct of the study. The study protocol was endorsed through rigorous scientific ethical review by the National Institute of Medical Research (NIMR) (ethical approval NIMR/HQ/R.8a/Vol. IX/444 granted on 27/10/2023). All participants signed a written informed consent prior to inclusion. All methods were performed in accordance with the relevant guidelines and regulations.

### Consent for publication

I hereby provide consent for the publication of the manuscript detailed above, including any accompanying images or data contained within the manuscript.

### Availability of data and materials

The datasets used and/or analysed during the current study are available from the corresponding author on reasonable request.

### Authors’ contributions

Principal investigator: AD; Study design: AD, JPAL, VM, PM; Data collection: AD, PM, VM; Transcription: VM, AD; Data analysis: PM, AD; wrote draft of manuscript: AD; and reviewed and approved the final draft of the manuscripts: JPAL, PM, AD, VM.

## Acknowledgments

We thank all patients, qualitative researchers, and medical staff at Tanga Regional Referral Hospital, Tanzania and the Institute of Medical Research for their assistance and continuous support. Group Authorship requested.

## Supporting Information

## Appendix A Voices of Experience: Thematic Categorization of Participant Testimonies on Health Challenges and Practices

*All names or identifying labels used in participant quotes are pseudonyms, indicated by quotation marks, to protect participant confidentiality. Any direct references to specific locations have been replaced with [Redacted].*

### Voices of Experience: Thematic Categorization of Participant Testimonies on Health Challenges & Practices

#### Delay in treatment-seeking

“I came late to the hospital, believing this was just a fungus…” (P1, Female)

“It took like six months without going to the hospital…” (P2, Male)

“Yes, I was late to go to the hospital, I think I should have been when I started scratching myself …” (P4, Female)

“Sometimes you reach a point where you give up because when you apply it, you see no results…it deprives you of strength …” (P2, Male).

“If it were, say, identified early, and I had gone to the hospital sooner, I wouldn’t have reached this stage. They (traditional healers) delayed me, yes…” (P2, Male)

“Unfortunately, it started as a fungus, so I thought it was a fungus. I treated with antifungal drugs because most of the time fungi appeared on the feet, at the end of the day, the leg became swollen and started to turn a little brown, then I knew there was a problem …” (P9, Male)

#### Wound Management

“When I go to shower, because it started here at the bottom, I clean it, and then I apply medicine, thinking it’s a fungus. It was negligence, thinking it’s just a fungus because I didn’t trip, wasn’t pricked by a thorn, or cut myself…When I noticed… uh… my foot was swelling, that’s when my children, who don’t live with me, got alarmed and took me to the hospital” (P1, Female).

“I used to go to the pharmacy and buy a sterile gauze, I clean it and then I apply the medicine knowing that it is just a fungus… I was covering my wound with a piece of cloth like this so as to protect myself from the sand and step on… believing that it won’t affect anything …[I used] home clothes, my child” (P1, Female).

“I pour iodine on the gauze and then clean the wound. After cleaning, I apply the same iodine. Just once a day, mainly when I take a shower” (P1, Female).

Ah, for the first month I cleaned it every day, but in the following month, I would skip a day and then go dress it, like dress it today, skip tomorrow, then go the day after. I was unable to go frequently because of the financial challenge…I had to pay five thousand for dressing and four thousand for transport costs to reach the health centre every day … (P4, Female).

“ … The lack of a cure for diabetes is a big problem, it’s humiliating, it means you get a toe amputated today, and a leg tomorrow” (P5, male).

“You never cleaned the wound yourself?” - (PM).

“Uh uh (disapproving)” (P3, Male).

“They [Muhimbili hospital] cleaned me well, gave me medicine, a lot of blood transfusion, it started forming flesh properly, and finally I was discharged [after four months], the skin had attached well, I stayed at home, but because I’m from [redacted], I went back home and after about four months, the condition started forming pus again, inside the leg, and the situation reappeared again. And it reappeared again, and that’s the condition I’m in now” (P2, Male).

“I cleaned [the wound] myself. I just cleaned it using water or sometimes I cleaned it using the same healer’s medicines, like if someone gives you water to mix, so I cleaned it then later there are other medicines he gives you to apply … once a day” (P2, Male).

“I was just washing it with metronidazole and dressing it with honey” (P4, Female).

“I continued using these ordinary medicines like paracetamol” (P10, Male).

#### Ritual Practices

“I visited like eight traditional healers … after using traditional medicine for three years … the condition became worse … you can apply medicine today, until tomorrow the leg will be sore and itchy …” (P2, Male).

“The second (traditional healer) said something like… I don’t know… there are … two birds with two beaks, that’s why you have sores here and there, that’s what’s troubling you. (Laughter) … the third traditional healer [said] it’s because of spirits… I don’t know, they want me to become a traditional healer, so I need to perform rituals to become a healer and that condition will go away..each one came with their own reason. He claimed…(long pause)… that I had stepped on, I don’t know, mermaid children on the road… at a crossroads …” (P2, Male).

“I used the bark of a tree called Mzambarao … I would feel less tired after drinking it, and my frequent urination reduced. [I drank it] twice a day, two small cups … I got that advice from another diabetic patient … effectiveness was … equivalent to the pills because it lowered the sugar level faster … The pills did lower it, I don’t deny, but their action was a bit slower” (P6, male).

“These Western medicines from the hospital didn’t work. Every time I take these hospital medicines I don’t get relief, I met a gentleman, he said let me give you medicine. He gave me a leaf and I chewed it and put it on my wound” (P9, Male).

“A belief in what? Superstition. That is, they don’t believe that the problem was caused by God but immediately see it as caused by another human. So they can’t solve it without resorting to superstition” (P1, Female).

“We used to drink herbal medicines a long time ago but not for wounds. Since I got burned, I’ve never taken herbal medicines” (P3, Male).

“Yeah, some healers tell you to use [the medicine] three times a day if it’s for drinking, and for ointments, there are some that you apply to the wound, you apply it. He might tell you to apply it today, then clean it tomorrow and reapply, or apply in the afternoon and then in the evening. The medicines are so strong, they itch and hurt all day” (P2, Male).

“Diabetes started for me in 2011 … unfortunately, we have this problem in my family. My younger brother told me, “Forget the hospital medications. Look for herbal remedies.” I started using those Chinese herbs, until 2016 I couldn’t find them anymore. I completely missed them. Then, fortunately, although I had used many medicines before and saw they didn’t work. Many turned out to be lies, someone tells you three hundred thousand TSh, then you leave them. I used many. But around 2016, I met an old man in the village, “Ali Magwadu”. That man has good medicine, really. For diabetes, eh, I appreciate it. He gave me these large capsules for a month, I could take them and stay well for a year. Without needing anything else” (P9, Male).

“He gives you roots. There are many types of roots. So he cuts them and gives you up to seven days’ worth. So you take it for a month or two. It’s a very good medicine. I really appreciate that it’s the medicine for diabetes, yes” (P9, Male).

#### Eating Behavior

“At night… we eat light foods like bananas … or cassava with tea” (P1, Female).

“I had never followed any diet, I eat ugali, rice and sometimes I miss the food completely … I eat meat but once in a while depending on the economic situation, if I get it today I might go a whole month without it again” (P2, Male).

“I often don’t eat at lunch” (P4, Female).

“In the morning, I can only drink porridge…And eat nothing until the next morning…I can’t taste food…But surprisingly, the sugar level doesn’t stabilize” (P5, Male).

“In the morning sometimes I might have tea, sometimes I might miss it, yes if I get it, in the afternoon I eat ugali, if I get rice I eat, I don’t have a specific diet, maybe this diet is because I haven’t got it for my health yet” (P2, Male).

“I usually eat boiled bananas…around 9 a.m and before I go to work, I must have a cup of this (indicating something) porridge. In the afternoon, I eat ugali. Sometimes at night, I eat potatoes, those kinds of boiled potatoes, yes” (P6, Male).

“First thing in the morning I drink millet porridge. In the afternoon, I cook dried bananas” (P10, Male).

#### Inadequate Healthcare Services

“No, [HCW’s did not give instructions on cleaning the wound] because they cleaned it themselves” (P1, Female).

“When I got to the hospital, they said let’s rest her because she is diabetic, let’s give her first aid, so I stayed from Saturday, Sunday, around Monday, Tuesday, then they said no, let’s take her to the city, to the regional hospital for an X-ray, to understand the problem and how it’s progressing” (P1, Female).

“Maybe just the biggest challenge is the delay in treatment. Since I arrived here on Tuesday evening, they have already set their treatment time table only for Tuesday and Wednesday of next week. So it bothered me and it was difficult to get treatment and I see that the bacteria are attacking more and more now …” (P9, Male).

“I went to the district hospital, I didn’t get any treatment there. I stayed there for four days, so I had to come back to the referral hospital …” (P10, Male).

“[The HCWs] tell us to eat good food, soup, small fish, spinach and vegetables” (P3, Male).

“At that time I just thought it was something ordinary that would heal even with local remedies. But after trying local treatments and failing, I started going to the health center. So even after reaching the health center, they would clean it, give me pills, injections, but it kept growing, worsening, and spreading over my leg” (P2, Male).

“I started feeling unwell about three months ago. It began as a blister, like I started itching, and then the blister burst and turned into a wound. The wound grew bigger and bigger, so I went to a private hospital. There, I was dressed and given medication, I would go home, get dressed again, and medicate, but… it seemed fine for a while, then it developed slough, so I had to come here” (P4, Female).

“[At the private hospital] I was using povidone, I was using… there are these drips, metronidazole (inaudible)… these small drips called metronidazole, for cleaning. it improved again but then deteriorated, like it developed some sort of fungus. [At the hospital] they did not give me any dietary instructions, only that after being dressed, when bathing, I should keep it dry. I wear a wrapper and elevate my leg while bathing to keep it dry. After a month I went to [redacted] Hospital, but … the wound was not healing well, that’s why I came to this hospital (TRRH)” (P4, Female).

“At the hospital, they dressed it with NS, I was cleaned with NS and then dressed with honey. Just once a day” (P4, Female).

“There’s a doctor there named “Niko”, we went… he was called to my home, he came and saw it and said it’s not a big deal, let’s just inject some drying injections. So, he injected me with five injections, but when they were finished, I still felt pain and the swelling increased. I [went to my] diabetes clinic at [redacted] hospital, when I got there, they looked at it and said the foot might have been damaged so they gave me a transfer to go to Bombo (TRRH) and they removed my toe” (P5, Male).

“After being discharged on the 29th, I was given instructions on using… (long pause). I was told that I must use oil on the dressed foot…After one week, I was told to dress it using honey, and I did that. It was a daily clinic. At Tanga Central” (P5, Male).

“They gave me a medicine called povidone iodine. They gave me pieces of gauze and bandages. They told me to clean it first with spirit, and after cleaning it with spirit, then dress it with povidone.[I did this] twice a day, in the morning, but I didn’t use spirit, I used warm water with salt” (P6, Male).

“The most important thing they [hospital] told me was to use honey. I have never used honey as a treatment, you see, it was something I didn’t believe in. They did not give me any dietary guidelines” (P6, Male).

“Yes, I went to the health center, they told me to come every day to get dressed. They were applying that povidone, iodine. Yeah, I was applying in two places. Every day I applied povidone. But unfortunately, the wound started smelling worse. I applied povidone, and it smelled worse. But I also applied that leaf. The leaf refused to stay. That’s the problem I had. Because there on the toe, I don’t know if it’s due to friction or what, the leaf just refused to stay. Every time I applied it, it fell off” (Patient 15)

#### Living and coping with chronic wounds

“Ah, challenges, truly… Ah, there aren’t any, just worries. Anxiety because you know, often with diabetic wounds, you hear about people having their legs amputated … you just feel the pain, it’s like pricking - Ah - it’s sharp. Yes, but when you step on it - Ah - no problem, and secondly, the fluid it releases has a bad smell, so when you sit with others… you find flies following you, you can’t sit comfortably” (P6, Male).

“The challenge was that my foot was swollen… it was badly swollen, so walking and wearing shoes or sandals was difficult” (P1, Female).

“I had a severe headache and I collapsed unconsciously. One leg was left outside, and the other was near the fire. So for about two hours, that leg was next to the fire. When I woke up, I was surprised to find one leg completely numb. When I touched it, my arm, neck, and side up to my neck were numb. They took me to Lushoto Hospital, our district hospital. When I got there, they tested me and said my blood pressure was high. They gave me medicine, and all the numbness subsided … When they saw the burn was severe, they referred me to Tanga, Bombo. [Redacted hospital] saw they couldn’t handle it because it’s a small hospital” (P3, Male).

“Then I came here, they took me and removed the dry flesh, I went to the theater, and they fixed an exposed bone. Yes, I’ve been treated, and now it’s the wound that’s bothering me. The main challenge with the wound is just the pain… it hurts, especially when being cleaned, until you take painkillers or get an injection, then it calms down” (P3, Male).

“I don’t have insurance. My situation is humble. My children contribute, and we pay” (P3, Male).

“I’m stuck, yes, like the other day I went to the theater, I went to get grafts, they refused to take. Yes, that’s why they came to tell me to take the fluids for research on the wound, maybe that is why it refused. Not that I’m refusing because I don’t have a specific job” (P3, Male).

“Sometimes when you apply some medicines, like if you apply it today, you can stay with it almost from today to tomorrow or the day after, and that leg still hurts and itches, it pulls and makes you uncomfortable, so challenges like that, I had no peace at all” (P2, Male).

“The challenge with the schedule was sometimes he might tell you to do this or he comes and you do this, sometimes you don’t follow exactly what he suggests [because] sometimes you reach a point where you give up because when you apply it, you see no results, that’s why you think maybe if someone else appears because maybe I should leave this one first, because there’s that challenge, it deprives you of strength, even the appetite to eat sometimes, you don’t eat because of the suffering you get from the pain of those medicines” (P2, Male).

“I’ve been given instructions, but the (stuttering) economic situation is challenging. I think it [treatment] can even reach three hundred thousand. Because they told me the first one, they said I don’t know one hundred and fifty thousand… one hundred I don’t know and… One hundred, I don’t know and bit.. Something like that, then the second one for scraping they said one hundred and fifty thousand. The plan here, I think maybe after I finish being given blood because I… I came in here because I was lacking blood, so I think maybe after I finish the blood transfusion, my schedule as advised, maybe I leave first maybe to look for… people to organize, if money is found then I come and get this issue handled” (P2, Male).

“I really believe if I had gone to the hospital earlier I wouldn’t be in this condition … I just believed it would heal in time, but it wasn’t as I thought … it’s not good to self-treat at home, because there are many things you don’t understand. It’s better to go to the hospital early to know the proper procedure-…the dangers are like this, the wound becomes big and you stay with it for a long time” (P4, Female).

“my body becomes very heavy, I can’t handle it, yes. I just can’t. For example, I might go outside, but it’s a challenge. I might fall … It feels like my legs are weakening, yes. And, you might use the toilet but then find it hard to get up. In short, I can’t manage on my own” (P5, Male).

“My opinion is that more effort should be made to find a cure for diabetes … Yes, the lack of a cure for diabetes is a big problem, it’s humiliating, it means you get a toe amputated today, a leg tomorrow … it will be a continuous process now. Removing these body parts. If this foot is problematic, are there no other treatments other than amputation?” (P5, Male).

“I told them, look, this is not enough, I need to move forward, because now it’s starting to produce fluid with a strong smell. I saw no progress staying there, why should I stay? What I did was just ask for a paper. Just give me a paper to go to Bombo. I feel like we’ve failed here” (P6, Male).

“I sometimes take a crepe bandage, it’s light, but I just wrap it lightly around, so I can sit with others inside there and talk to them without flies bothering me” (P6, Male).

“If there was a treatment, people wouldn’t have limbs amputated, there wouldn’t be removal of body parts, but now it has become a critical issue. Yes, sir. An issue that is now being talked about a lot, eh. And also, people have continued to be… You know, a disease is not rejected, it’s accepted because it has already reached you, but now people are scared of it, eh, they are afraid. So, I continue to pray that you continue researching so we can find medicines that can cure these wounds” (P6, Male).

“These sugar testing screens, they are expensive. But without testing you can never know. And when you come to the hospital you are told there are no sticks, no this or that. So it’s better to have your own” (P9, Male).

“Ah, I don’t really understand the medications I take” (P10, Male).

#### Suggestions from patients

“My opinion to my fellow Tanzanians is that when we get a minor wound, we shouldn’t ignore it but rush to the hospital” (P1, Female).

“Ah, my suggestion, which I had, first is to thank them, to thank them for continuing to try to research, yes, in order to help more diabetic patients like us, especially when we get wounds like these, if possible to find a treatment” (P6, Male).

“My opinion is that more effort should be made to find a cure for diabetes … the lack of a cure for diabetes is a big problem, it’s humiliating, it means you get a toe amputated today, a leg tomorrow … it will be a continuous process now. Removing these body parts … people aren’t delving deeply enough technically because if this foot is problematic, are there no other treatments other than amputation? That’s my question” (P5, Male).

“Uh, other recommendations, it’s not good to self-treat at home, because there are many things you don’t understand. It’s better to go to the hospital early to know the proper procedure- the dangers are like this, the wound becomes big and you stay with it for a long time” (P4, Female).

“Ah, we thank you for your attention. You’ve come to listen to us, I believe even this is treatment for us… we learn…I know even you when you get your research I’ll have been helped. Suggestions maybe just continue helping us, once you get your research come help us..help us so we can go back to the streets … I just pray God helps you continue being this way” (P2, Male).

## List of abbreviations

AMR: Antimicrobial Resistance
HCW: Healthcare Workers
IDI: In-depth Interviews
IRB: Institutional Review Board
KAP: Knowledge, Attitude, and Practices
MDR: Multidrug resistance
MTA: Material Transfer Agreement
NatHREC: National Health Research Ethics Committee
NIMR: National Institute for Medical Research
OTC: Over the counter
PI: Principal Investigator
REIMS: Research Ethics Information Management System
SSA: Sub-Saharan Africa
TRRH: Tanga Regional Referral Hospital

## References

[1] Thorud JC, Plemmons B, Buckley CJ, Shibuya N, Jupiter DC. Mortality after nontraumatic major amputation among patients with diabetes and peripheral vascular disease: a systematic review. J Foot Ankle Surg. 2016;55(3):591–599. doi:10.1053/j.jfas.2016.01.012

[2] World Diabetes Foundation. Diabetes foot care - Step-by-Step. Bagsværd, Denmark: World Diabetes Foundation; Available from: https://worlddiabetesfoundation.org/what-we-do/projects/wdf03-0056/

[3] European Commission. Special Eurobarometer 407: Antimicrobial resistance [Internet]. Brussels: European Commission; 2013. Available from: https://health.ec.europa.eu/system/files/2016-11/ebs_407_sum_en_0.pdf

[4] Kaanto R, Kagira J, Waititu K, Ngotho M, Maina N, Gachohi J. Characteristics of patients presenting with septic wounds in selected hospitals in Kajiado County, Kenya. Int J Community Med Public Health [Internet]. 2021 Jul. 27;8(8):3743–9. Available from: https://www.ijcmph.com/index.php/ijcmph/article/view/8245

[5] Pieper B, Sieggreen M, Nordstrom CK, Freeland B, Kulwicki P, Frattaroli M, et al. Discharge knowledge and concerns of patients going home with a wound. Journal of Wound, Ostomy, and Continence Nursing: Official Publication of The Wound, Ostomy and Continence Nurses Society [Internet]. 2007 May 1;34(3):245–53; quiz 254-255. Available from: https://pubmed.ncbi.nlm.nih.gov/17505242/

[6] Dawer A, Msengi V, Mtui TB, Sarakikya S, Suleiman R, Lusingu JPA. Antimicrobial resistance of bacterial isolates among patients with chronic wound infections in Tanga Regional Referral Hospital, Tanzania. Unpublished manuscript. 2025.

[7] Global Data Lab. Health indicators. Nijmegen (NL): Global Data Lab, Institute for Management Research, Radboud University. Available from: https://globaldatalab.org/health/, version v1.0.

[8] Gould L, Abadir P, Brem H, Carter M, Conner-Kerr T, Davidson J, et al. Chronic wound repair and healing in older adults: current status and future research. Wound Repair Regen. 2015;23(1):1–13. doi:10.1111/wrr.12245. PMID: 25486905; PMCID: PMC4414710.

[9] Rizvi MK, Rabail R, Munir S, Inam-Ur-Raheem M, Qayyum MMN, Kieliszek M, Hassoun A, Aadil RM. Astounding health benefits of Jamun (*Syzygium cumini*) toward metabolic syndrome. Molecules. 2022 Oct 24;27(21):7184. doi:10.3390/molecules27217184. PMID: 36364010; PMCID: PMC9654918.

[10] Alhassan AR, Kuugbee ED, Der EM. Surgical healthcare workers’ knowledge and attitude on infection prevention and control: a case of Tamale Teaching Hospital, Ghana. Can J Infect Dis Med Microbiol. 2021;2021:6619768. doi:10.1155/2021/6619768. PMID: 33981370; PMCID: PMC8088377.

[11] Woo KY, Wong J, Rice K, Coelho S, Haratsidis E, Teague L, Rac VE, Krahn M. Patients’ and clinicians’ experiences of wound care in Canada: a descriptive qualitative study. J Wound Care. 2017 Jul 1;26(Sup7):S4–S13. doi: 10.12968/jowc.2017.26.Sup7.S4. PMID: 28704169.

[12] Dorai AA. Wound care with traditional, complementary and alternative medicine. Indian J Plast Surg 2012;45:418–4.

[13] Asuzu CC, Akin-Odanye EO, Asuzu MC, Holland J. A socio-cultural study of traditional healers’ role in African health care. Infectious Agents and Cancer. 2019 Jun 20;14(1).

[14] Albertyn R, Berg A, Numanoglu A, Rode H. Traditional burn care in sub-Saharan Africa: A long history with wide acceptance. Burns. 2015 Mar;41(2):203–11.

[15] Tyavambiza C, Dube P, Goboza M, Meyer S, Madiehe AM, Meyer M. Wound Healing Activities and Potential of Selected African Medicinal Plants and Their Synthesized Biogenic Nanoparticles. Plants. 2021 Nov 30;10(12):2635.

[16] Taber JM, Leyva B, Persoskie A. Why do people avoid medical care? A qualitative study using national data. J Gen Intern Med. 2015 Mar;30(3):290–7. doi:10.1007/s11606-014-3089-1.

[17] Jan M, Almutairi KH, Aldugman MA, Althomali RN, Almujary FM, Abu Mughaedh NA, Alhadi LN. Knowledge, attitudes, and practices regarding wound care among general population in Aseer region. J Family Med Prim Care. 2021 Apr;10(4):1731–1736. doi:10.4103/jfmpc.jfmpc_2331_20. PMID: 34123920; PMCID: PMC8144783.

[18] Van Wyk B-E, Moteetee NA. Ethnobotanical research in sub-Saharan Africa – documenting and analysing indigenous knowledge about medicinal, edible and other useful plants. S Afr J Bot. 2019 May;122:1–2. doi:10.1016/j.sajb.2019.04.020.

[19] Priyadarshani RAC, Samarawickrama MB. Knowledge, attitudes and practices of patients regarding chronic wound care and point prevalence of chronic wounds at surgical and medical units in Teaching Hospital Karapitiya, Sri Lanka. IOSR J Pharm Biol Sci. 2017 Mar–Apr;12(2 Suppl 2):38–46. Available from: https://www.iosrjournals.org/iosr-jpbs/pages/12(2)Version-2.html

[20] Bejarano-Roncancio, Jhon Jairo, Eugenia M, Saurith-López V, Sussman-Peña OA. Revista de la Facultad de Medicina [Internet]. 2025; 59:3–11. Available from: http://www.scielo.org.co/scielo.php?pid=S0120-00112011000500002&script=sci_abstract&tlng=pt

[21] Van Hecke A, Grypdonck M, Defloor T. A review of why patients with leg ulcers do not adhere to treatment. Journal of Clinical Nursing. 2009 Jan 14;18(3):337–49.

